# Human and computer attention in assessing genetic conditions

**DOI:** 10.1101/2023.07.26.23293119

**Authors:** Dat Duong, Anna Rose Johny, Suzanna Ledgister Hanchard, Chris Fortney, Fabio Hellmann, Ping Hu, Behnam Javanmardi, Shahida Moosa, Tanviben Patel, Susan Persky, Ömer Sümer, Cedrik Tekendo-Ngongang, Tzung-Chien Hsieh, Rebekah L. Waikel, Elisabeth André, Peter Krawitz, Benjamin D. Solomon

**Author notes:** Correspondence: Benjamin D. Solomon, *National Human Genome Research Institute*, Building 10 - CRC, Suite 3-2551, 10 Center Drive, Bethesda, MD 20892. Equal contributions (Dat Duong, Anna Rose Johny, Suzanna Ledgister Hanchard, Chris Fortney).

## Abstract

Deep learning (DL) and other types of artificial intelligence (AI) are increasingly used in many biomedical areas, including genetics. One frequent use in medical genetics involves evaluating images of people with potential genetic conditions to help with diagnosis. A central question involves better understanding how AI classifiers assess images compared to humans. To explore this, we performed eye-tracking analyses of geneticist clinicians and non-clinicians. We compared results to DL-based saliency maps. We found that human visual attention when assessing images differs greatly from the parts of images weighted by the DL model. Further, individuals tend to have a specific pattern of image inspection, and clinicians demonstrate different visual attention patterns than non-clinicians.

## Introduction

Deep learning (DL) and other types of artificial intelligence (AI) are extensively and increasingly employed in many biomedical areas, including genetics. In the field of genetics, AI is used for base calling from genomic sequencing, genetic variant classification, clinical note processing, and to help assess radiologic data.^1^ Among these applications, AI-based tools that can generate differential diagnoses based on facial images of individuals with possible genetic conditions have become widespread in both research and clinical contexts.^2,3^

Despite controversies and unsettled questions, healthcare is poised for major near-term changes as AI is implemented into workflows.^4^ One crucial issue involves understanding how AI tools perform and how “choices” made by AI compare with those made by humans, including people with different levels of expertise.^5-7^ This can be helpful to test the accuracy of models and to examine whether confounders are present. For example, an analysis of DL to assess chest X-rays for signs of COVID-19 showed that classifiers can be highly accurate compared to humans but may rely heavily on confounding information, such as radiographic clues outside of the lung fields.^8^ As another example, large-language models have shown extraordinary recent progress; while these models perform well compared to humans, they may present coherent but incorrect or inconsistent information.^9^

To explore these issues further in the context of genetics, we conducted an experiment of visual attention on images of individuals with or without genetic conditions. We approximate how well the visual attention maps of medical experts align with the saliency maps of a neural network model. We also examine and compare the visual activity of geneticist clinicians and non-clinicians. We find that saliency maps differ substantially from regions of human attention and that clinicians and non-clinicians assess images differently.

## Methods

### Data collection

Similar to our previous methods,^10,11^ we selected publicly available images of individuals between 2 and 18 years of age affected by one of 10 genetic conditions: 22q11.2 deletion syndrome (22q11DS), Beckwith-Wiedemann syndrome (BWS), Cornelia de Lange syndrome (CdLS), Down Syndrome (DS), Kabuki syndrome (KS), Noonan syndrome (NS), Prader-Willi syndrome (PWS), Rubinstein-Taybi syndrome (RSTS1), Wolf-Hirschhorn syndrome (WHS), and Williams syndrome (WS), as well as images of unaffected individuals similar in age and ancestral diversity to the affected individuals. The genetic conditions were chosen as they represent relatively common genetic conditions that involve recognizable craniofacial features, and with which geneticists would be expected to be at least somewhat familiar.^12^ Two geneticists (BDS, CTN) and a genetic counselor (RLW) on the study team selected the images to represent typical images representing the chosen conditions; images of some conditions were felt to be more difficult to recognize than others. Sixteen images (one image for each of the above conditions, and six images of unaffected individuals) were selected for the eye-tracking experiments. The selected images were at least 720-pixel resolution; they were each standardly cropped, centered, and aligned (e.g., the eyes, nose, and mouth were roughly repositioned at the same coordinates for all images). In our paper to, we include mentions of the figures we prepared; as preprint servers may understandably not allow such images to be shown, even when previously published, we include an asterisk (*) by each figure number to explain why it is not available in this version, or why a portion of the figure is not shown. See **Supplementary Figure 1\*** for image sources; images are shown as the standardized, processed versions used in the eye-tracking experiments. Note that all images shown are versions of previously published and/or publicly available images.

### Eye tracking experiments

The formatted images were embedded in a screen-based eye-tracking system (Tobii Pro X3-120, Tobii Lab Pro version 1.194.41215; https://www.tobii.com/, Stockholm Sweden). Eye-tracking experiments took place in two locations: the National Institutes of Health (NIH) (Bethesda, Maryland, United States) and the University of Bonn (Bonn, Germany). The study was considered exempt from formal IRB review. After calibration, each participant viewed the 16 images for seven seconds per image, and answered questions about each image, including whether the image showed a person affected by a genetic condition, and, if so, what condition the person had. After extensive initial testing, we chose seven seconds for the viewing time, as subjective feedback and preliminary assessments showed that this amount of time was sufficient to assess an image but minimized visually revisiting areas of the image in a way that might not further inform the assessment. To minimize head movement or distractions, questions were asked verbally during the eye-tracking portions of the experiment. Responses were documented manually by a study team member.

The NIH cohort included 17 individuals, including physician geneticists (14) and physicians in genetics subspecialty training (3). The Bonn cohort included 29 total individuals, including physician geneticists (2) and physicians in genetics subspecialty training (5), as well as 22 non-clinicians. Some of the non-clinicians in our cohort have some experience through their work in recognizing genetic conditions. For example, some are graduate students who study applications of AI in genetics, and thus are familiar with genetic conditions, but are not trained clinicians.

### Data extraction and analysis

We extracted eye-tracking data for analysis in two main ways. First, we extracted individual heat maps for each participant and image. We used the default Tobii software settings, but with two changes to enable our analyses: a) we changed the output eye-tracking radius to 25 pixels, as our preliminary analyses showed that the default 50 pixel radius was insufficiently precise for comparisons; b) we used a more homogeneous color scheme for the heat maps (with the following hexadecimal color codes: high: #943126, medium: #B03A2E, low: #CB4335), which enabled our quantitative analyses to take into account all captured heat map data, including to compare eye-tracking data with classifier saliency maps.

Second, to enable additional analyses that took into account gaze trajectory behavior to analyze the timing of participant gaze,^13^ a dysmorphologist (BDS) manually drew areas-of-interest (AOIs) for each image (see **Supplementary Figures 2a-j\***). The AOI set included only those features that were listed as having dysmorphic manifestations based on the clinical synopses in OMIM (https://www.omim.org/) and which were also present in the images. For example, if a condition were listed in the clinical synopsis section of OMIM as having a dysmorphic manifestation affecting the eyebrows and that manifestation was present in the image of a person with that condition used in the survey, an AOI was drawn around the eyebrows. Using the Tobii software, we extracted tabular data for analysis based on these defined AOIs.

Prior to analysis, we manually reviewed results and excluded heat maps where eye-tracking data was not recorded (this occurred for all results for one NIH participant, which may have been due to ophthalmologic issues like severe myopia, and seven total other isolated eye-tracking results for unclear reasons).

We observed that standard heat maps primarily showed that the areas around the eyes, nose, and mouth are the regions with the highest attention. These common visual attentions (**Supplementary Figure 3**), which likely reflect standard human behavior, make it difficult to quantitatively compare the visual activity differences between groups unless accounted for. To mitigate this issue, we computed the average heat map for all the clinicians and non-clinicians (separately) over all of the images. We then subtracted this common average gaze pattern (separately for clinicians and non-clinicians) from each heat map. This does not cause us to ignore these areas of common facial attention in our analyses but helps account for typical human behavior when viewing faces. **Figure 1\*** explains our data pre-processing approach.

**Figure 1.**
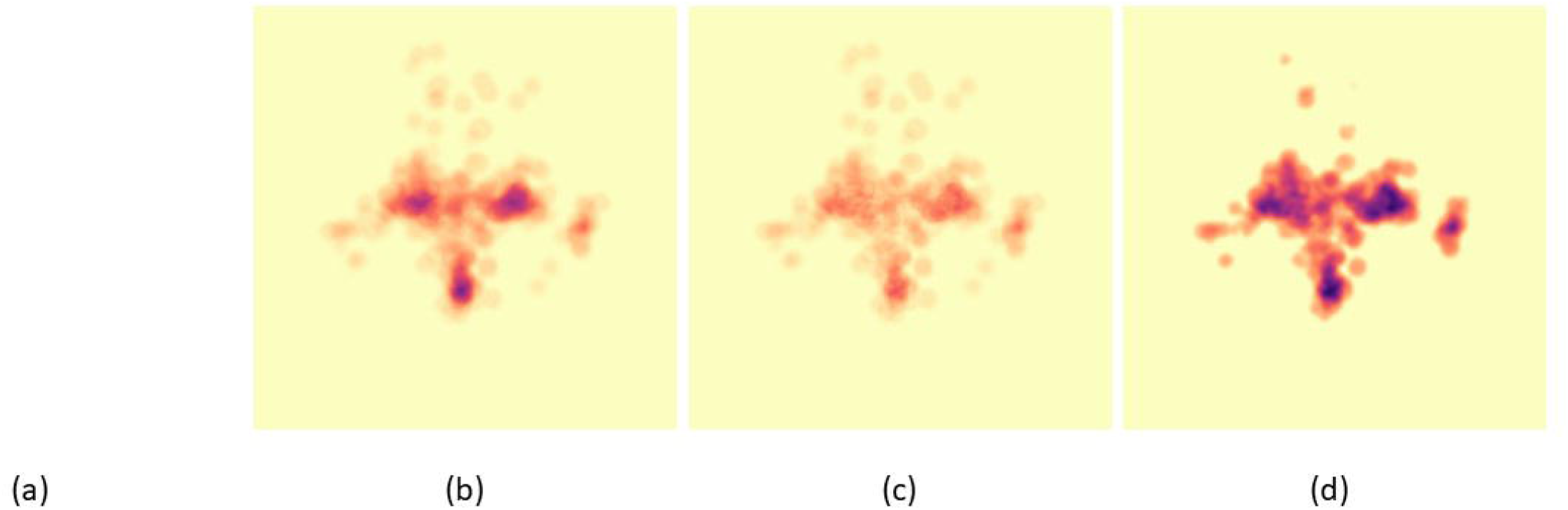
An example illustrating how we pre-process Tobii visual heat maps for subsequent analyses. (a) Original image of an individual with Kabuki syndrome (KS) (not shown due to image restrictions for this preprint server). {b) For this KS image, conditioned on the group of successful clinicians, we average the default attention heat maps. (c) We removed the common visual signals (See **Supplementary Figure 3)**. (d) Finally, we smooth the image with cv2.boxFilter and increase the color intensity. Here, comparing to (b), in (d) we can better observe the high visual interest at the lower part of the left eye in KS. **ALL** IMAGES OF INDIVIDUALS ARE PREVIOUSLY PUBLISHED AND/OR ARE PUBLICLY AVAILABLE - SEE SUPPLEMENTARY FIGURE 1 FOR REFERENCES FOR EACH IMAGE USED.

In our analyses of the images, we focused on four subgroups: clinicians and non-clinicians who correctly or incorrectly identified that an image represents a person with a genetic condition (e.g., see the categories in **Table 1**). One interest is whether the untrained intuition of a non-clinician aligns with clinician behavior. In the later sections of our analyses, we use the terms successful and underperforming clinicians and non-clinicians to refer to the clinicians and non-clinicians who correctly and incorrectly recognize the presence of a genetic disease for a given test image, respectively. For two different test images, the same participant may fail to recognize the disorders in both images. Hence, for two different images, the groups of successful clinicians (and likewise non-clinicians) may not have the same participants.

**Table 1.**
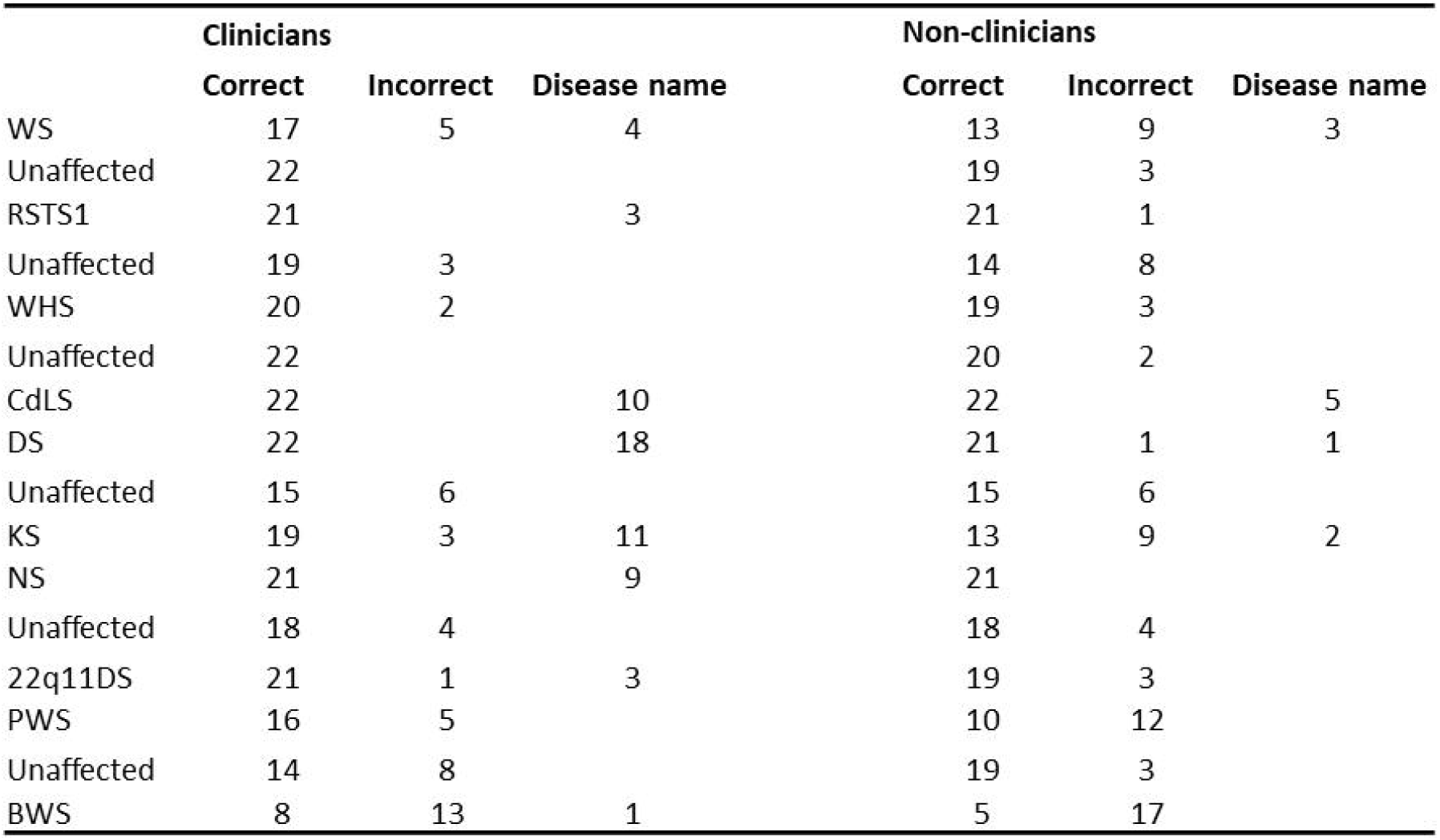
For a specific image, we count the number of clinicians and non clinicians who correctly and incorrectly identified that this image is of an affected or unaffected individual. Column *Disease name* indicates the number of clinicians and non-clinicians who correctly identified the disease name. As seven individual results had to be discarded for technical reasons, not every image has a complete data from 22 clinicians and 22 non-clinicians. The images are listed in the order shown to the participants.

|  | Clinicians |  |  | Non-clinicians |  |  |
| --- | --- | --- | --- | --- | --- | --- |
|  | Correct | Incorrect | Disease name | Correct | Incorrect | Disease name |
| WS | 17 | 5 | 4 | 13 | 9 | 3 |
| Unaffected | 22 |  |  | 19 | 3 |  |
| RSTS1 | 21 |  | 3 | 21 | 1 |  |
| Unaffected | 19 | 3 |  | 14 | 8 |  |
| WHS | 20 | 2 |  | 19 | 3 |  |
| Unaffected | 22 |  |  | 20 | 2 |  |
| CdLS | 22 |  | 10 | 22 |  | 5 |
| DS | 22 |  | 18 | 21 | 1 | 1 |
| Unaffected | 15 | 6 |  | 15 | 6 |  |
| KS | 19 | 3 | 11 | 13 | 9 | 2 |
| NS | 21 |  | 9 | 21 |  |  |
| Unaffected | 18 | 4 |  | 18 | 4 |  |
| 22q11DS | 21 | 1 | 3 | 19 | 3 |  |
| PWS | 16 | 5 |  | 10 | 12 |  |
| Unaffected | 14 | 8 |  | 19 | 3 |  |
| BWS | 8 | 13 | 1 | 5 | 17 |  |

To understand the general behavior of a specific participant group (e.g., successful clinician group), after accounting for the areas of common visual attention, we computed the average heat map for each test image over all the participants. Next, we applied two different thresholds to binarize an average heat map so that the values in this heat map are either 0 or 1.^14^ The first, lower threshold is meant to remove spurious noise. The second, higher threshold removes a large proportion of the signal; here, we would analyze only facial regions with the highest visual attention. At each of these thresholds, we use the intersection-over-union metric (IoU) to compare average heat maps.^14^ Standard deviation for the observed IoU was computed via bootstrapping. Finally, we applied random effects meta-analysis (we refer to this as “RE Model” in the Figures) to summarize the behavioral differences over many images between the two groups of participants (e.g., successful clinicians versus successful non-clinicians).

### Classifier and saliency maps

Our test set is the 16 images selected in the eye-tracking experiment. With the remaining images of the images collected (but which were not used for eye-tracking experiments), we trained EfficientNet-B4 using 5-fold cross-validation to build an ensemble model.^15^ Each image was resized to 448 × 448 pixels, and cross-entropy loss with equal weights for the diseases was used. Training process follows standard convention; for example, we tried different values for batch size and dropout rate and found that batch size 64 and dropout rate 0.2 worked best. We also found that a learning rate 0.00003 and the Adam optimizer were most effective for our analyses.^16^ For a test image, to combine the prediction of the models from 5-fold cross-validation, we average the classifiers’ outputs for this image.

The occlusion method with box size 20 × 20 pixels and stride 10 × 10 was used to produce the saliency maps.^17^ The intuition is to remove a 20 × 20 pixel box from the image and then measure how much the prediction accuracy drops. This idea allows us to identify key regions affecting the classifier’s output. We average the saliency map produced by the models from 5-fold cross-validation for each test image. We then apply a low and a high threshold to respectively remove just spurious signals and retain only the most important regions. Finally, we compute the IoU score for each image comparing the participant heat maps and the saliency map at the same filtering threshold (e.g., low or high for both human and model). We acknowledge that there are other models besides EfficientNet-B4 that have been studied in the context of genetic and other congenital diseases.^2,3^ Moreover, there are other saliency approaches besides the occlusion method. This paper’s combination of EfficientNet-B4 and the occlusion method seems to return reasonable results. In future studies, we plan to evaluate other types of image classifiers and saliency approaches.

Code is available at: 32.3
https://github.com/datduong/tobii-eye-track-syndromic-faces and https://github.com/datduong/classify-syndromic-faces.

## Results

### General accuracy and results

We divided the human participants into two main groups: 22 clinicians (15 from NIH and seven from Bonn) and 22 non-clinicians. **Table 1** compares the accuracy of the two groups at identifying whether an image shows a person affected or unaffected with a genetic condition. Overall, the clinician group was more accurate than the non-clinician group at recognizing whether an image showed an affected person (correct identification was 85.6% for clinicians versus 76.9% for non-clinicians, p = 0.0032 by Chi-square). In further responding to a question about what specific genetic condition the image might depict, non-clinicians were, unsurprisingly, usually unable to identify the disease names. However, some non-clinicians are familiar with specific conditions based on their work, such as involvement in genetic research.

Although our neural net (NN) classifier model was trained to predict 11 different labels (10 diseases and unaffected) for the conditions included in this study, when looking at unaffected and affected as the only two label choices, our classifier achieves 100% accuracy (see more details below).

### Evaluating human visual attention versus saliency map

NN models perform at least as well as humans at many image classification tasks, including the identification of genetic and other congenital diseases from facial images.^2,9,11^ For a specific input image, one can extract the corresponding saliency map from a NN image classifier. This saliency map indicates the regions of the image that the NN model deems to be relevant for the prediction of the ground truth label. Our NN classifier correctly labeled 13 out of the 16 test images with the specific genetic condition that person has; averaging over these 13 images, the prediction probability of the ground truth label is 0.7853. The three misclassified affected images are CdLS, PWS, and RSTS1. For these three images, the predicted probabilities of the ground truth label and incorrect label were: CdLS (0.3196 vs 0.3285 misclassified as NS), PWS (0.3383 vs 0.4690 misclassified as WS), and RSTS1 (0.0005 vs 0.7976 misclassified as NS). Because the classification accuracy of RSTS1 is low, we excluded this image in the analysis of this section (**Supplementary Figure 4** contains the analysis for all the affected images). We extracted the saliency maps for our NN classifiers to observe regions of the images that affect the model’s predicted probability for their ground truth labels. In other words, this approach allowed us to create saliency maps that are specific to each condition.

In this section, we evaluate whether the clinicians’ visual attention heat maps align well with a NN classifier’s saliency maps. We do not imply that humans and computers are doing the same thing when classifying an image, but this method allows a means of comparison. For example, a heat map shows how a single human participant assesses a single image. Although we can combine heat maps from different participants or of different images, generalizations and comparisons to the saliency map based on a NN may be difficult.

For the analyses, we treat the saliency maps as if they were heat maps, and then find the most reasonable threshold to binarize the saliency maps. Next, for a specific test image of an affected individual, we compute the IoU between the saliency map and the average heat maps of successful clinicians; examples are shown in **Figure 2\*** (see also **Supplementary Figures 5a-j\***).

**Figure 2.**
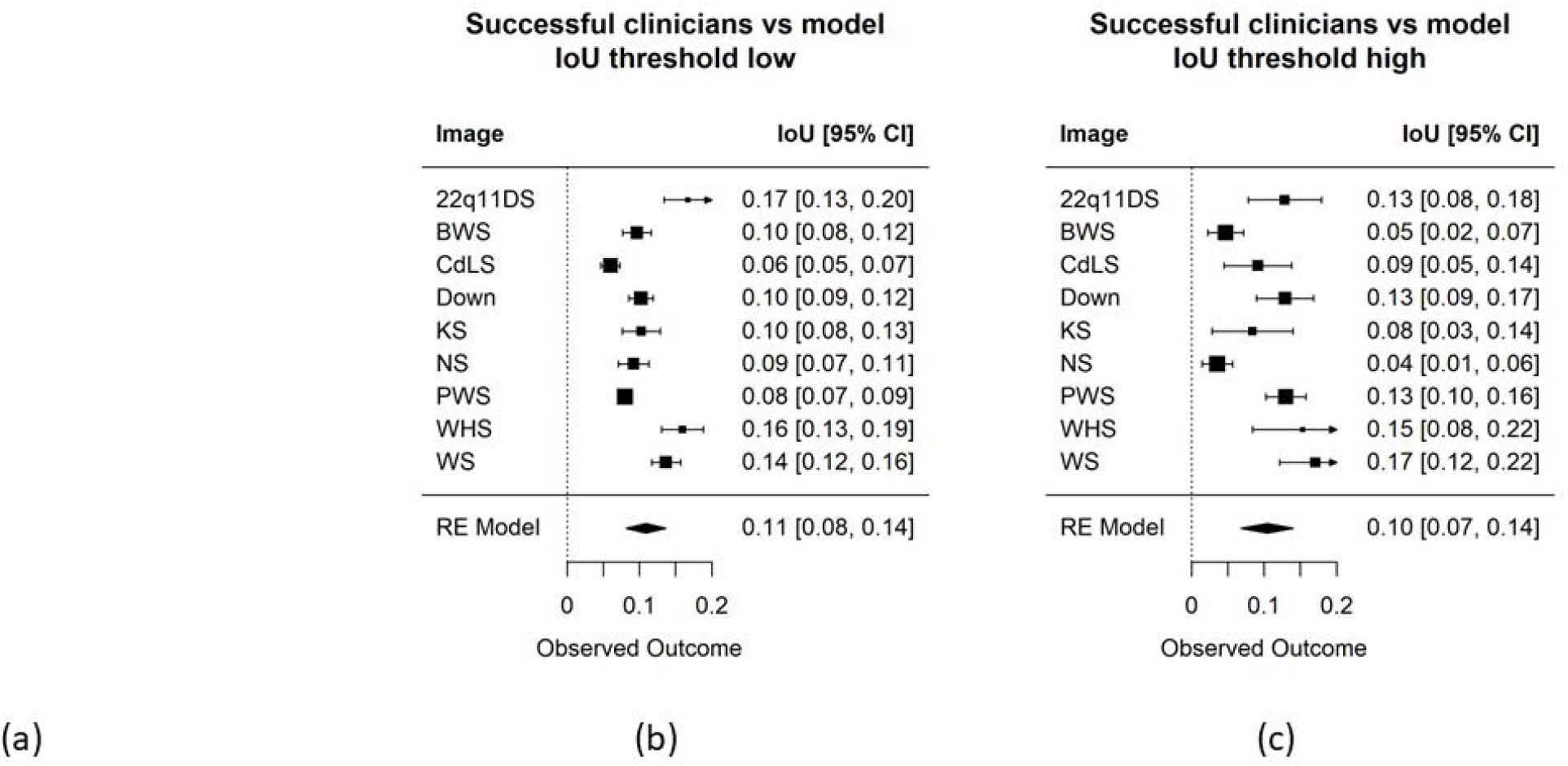
(a) Saliency maps of 22q11DS (top) and KS (bottom) test image highlight key regions that affect the classifier accuracy. Like the heat maps generated through eye-tracking experiments, we also apply a low (left) and high threshold (right) to the saliency maps. Visually, there are differences compared to the visual attention of human clinicians (not shown due to image restrictions for this preprint server). {b-c) loU metric compares the visual attention of successful clinicians and model saliency maps over the 9 test diseases (removing RSTS1) at the same filtering threshold (e.g., low and then high thresholds for both saliency maps and Tobii heat maps). ALL IMAGES OF INDIVIDUALS ARE PREVIOUSLY PUBLISHED AND/OR ARE PUBLICLY AVAILABLE - SEE SUPPLEMENTARY FIGURE 1 FOR REFERENCES FOR EACH IMAGE USED.

**Figure 2\*** also shows the IoU metrics averaged over the 9 affected images (excluding RSTS1 for reason mentioned above). We observed significant differences between human visual attention and what a NN model considers important. For example, the average IoU metric for successful clinicians versus the saliency map we generated was 0.11 for both low and high threshold heat maps. This IoU metric is much less than the metric comparing our clinicians and non-clinician groups (see below). In other words, human heat maps show that our participants tended to look at very different regions than the classifier per the models we examined.

### Analyses comparing groups of human participants

Using the methods described above, we compared the successful clinicians and non-clinicians for each of the 16 test images, and then summarized the visual attention differences over all these images. In **Figure 3\***, qualitatively speaking, the visual attentions of these clinicians and non-clinicians are more similar at a lower threshold, where only spurious visual signals were removed, and later become more different at a higher threshold, where only regions with high visual interests were retained (**Figure 3\***). Thus, as shown in **Figure 3\***, although these clinicians and non-clinicians correctly recognize affected versus unaffected individuals, on average these two participant groups do not show similar visual attention to the same facial regions; the IoU is 0.47 and 0.34 for low and high thresholds, respectively.

Figure 4. quantitively compares the visual interests of successful clinicians versus successful non-clinicians. Here, the result aligns with our qualitative inspection in **Figure 3**. When excluding low spurious signals, these two subgroups show similar eye-gaze interests (IoU 0.47). However, these two subgroups become more different when considering only the greatest visual attention (the IoU dropped to 0.34). This same trend is observed when we analyzed on just the set of affected and unaffected images (**Supplementary Figure 6**).

**Figure 4.**
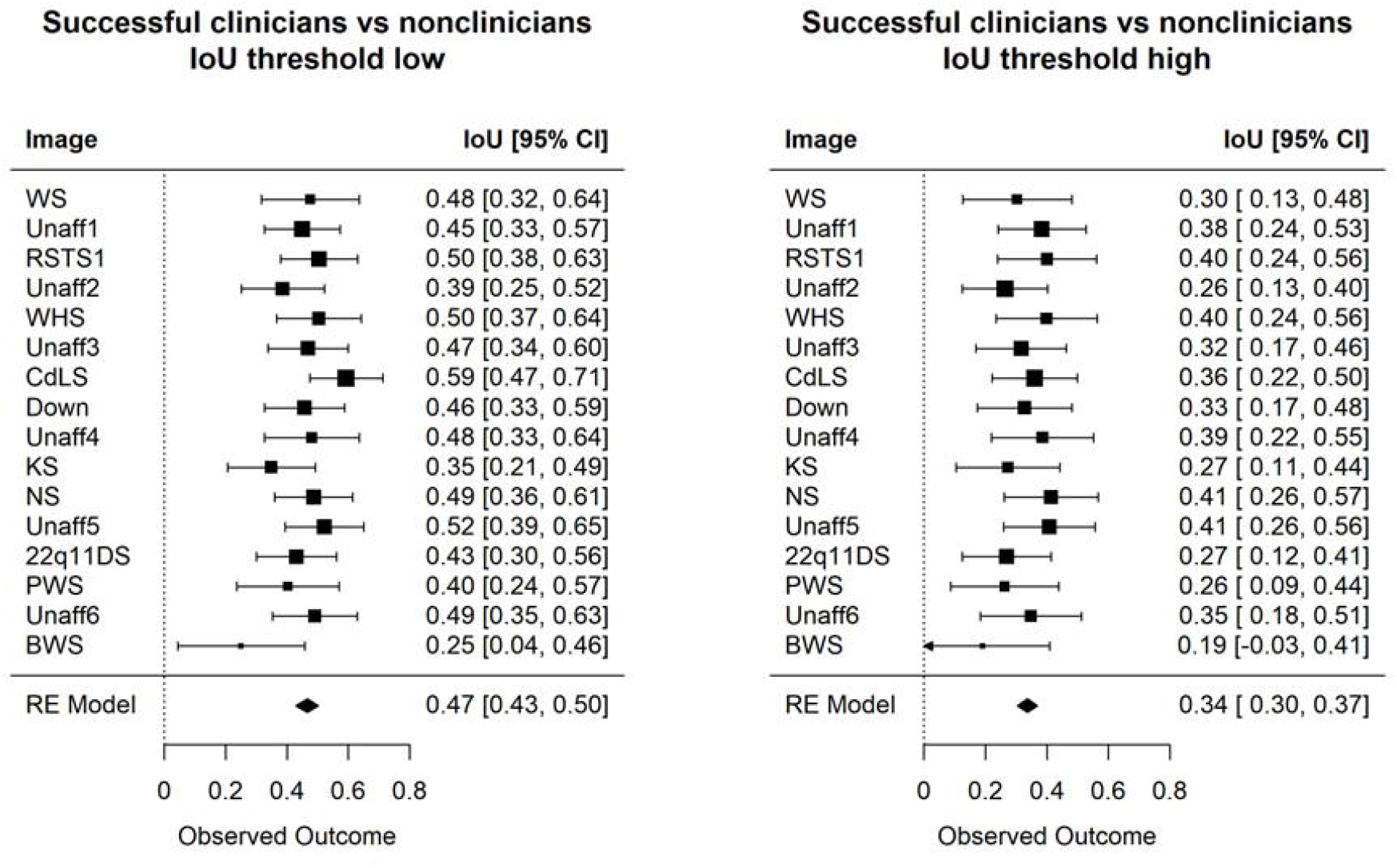
loU metric compares the heat maps of successful clinicians versus non-clinicians. When removing only low spurious signal (left), the two groups display more similar visual interests. However, when looking at only regions with high visual interests (right), the two groups differ more drastically. The images are listed in the order shown to the participants.

We further compare successful clinicians and underperforming clinicians. **Figure 5a** shows that, on average, these two subgroups of clinicians do not have similar visual interests. The same trend is seen when stratifying the non-clinicians according to their accuracy (**Figure 5b**). Conditioned only on participants who misclassified the images, **Figure 5c** also shows visual attention differences between clinicians and non-clinicians. Hence, when participants misclassify images, they have distinct ways of inspecting the images that do not appear to be similarly influenced by some common confounders in the images (e.g., hairstyles, facial expressions, or earrings that could affect a participant’s decision). Overall, there seems to be more similar visual attention when a participant (whether a clinician or not) can correctly identify that a person is affected by a genetic condition; for example, the IoU values in **Figure 4** are higher than those in **Figure 5**.

**Figure 5.**
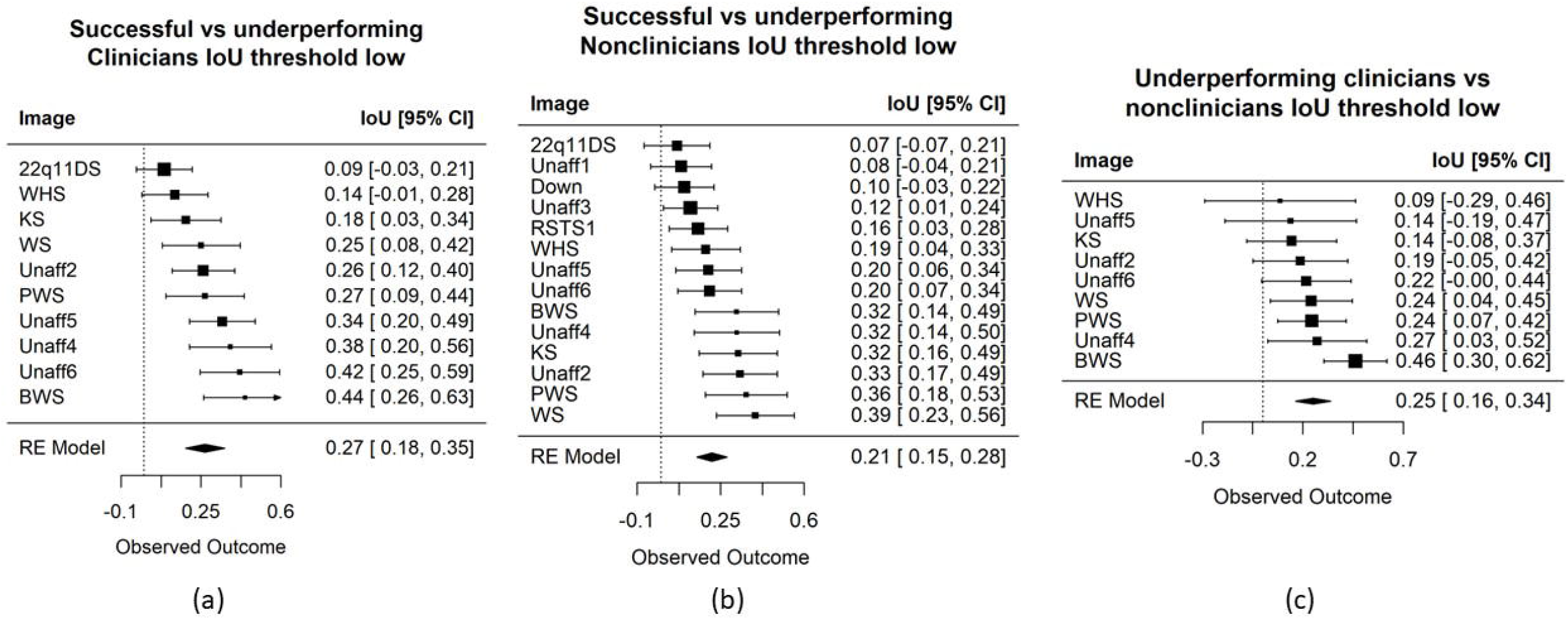
Images are sorted based on the loU metric, which compares the heat maps between different groups of participants. Here, we applied low filtering threshold to remove only low spurious signal from the average heat map of each group. These three comparisons show lower similarity scores than that of successful clinicians versus non-clinicians. Left: successful versus underperforming clinicians. Middle: successful versus underperforming non clinicians. Clinicians tend to show more similar visual interests for PWS, BWS, and unaffected images. This trend is not seen for non-clinicians. Right: underperforming clinicians versus non-clinicians.

We also examined the heat maps from the clinicians and non-clinicians, including to help investigate whether there were different visual patterns when clinicians examine affected or unaffected images. As shown in **Supplementary Figures 7 and 8**, participants had different individual patterns of assessing images. However, this individual pattern appeared consistent whether the participant observed affected versus unaffected images (see also **Supplementary Figure 9\***).

Finally, we examined the AOIs that corresponded to specific dysmorphic features in the images of affected individuals (**Supplementary Figure 2a-j\***). As each test image has more than one AOI that may be important to the underlying conditions, metrics like duration-of-fixation and time-to-first-whole-fixation for any single AOI can have high standard deviations, making it difficult to observe statistical significance. Overall, we did not detect a specific pattern for these AOIs that differentiated the categories of participants.

## Discussion

This study has two key results. First, human and computer attention differs substantially when evaluating images of individuals with potential genetic conditions. This does not mean that one is superior to the other, but these types of analyses and metrics may be helpful in future studies. For example, methods to compare human and computer attention can be used to explore potential confounders in AI-based analyses, or to develop methods that improve the accuracy or applicability of AI tools. As AI is increasingly adopted in clinical scenarios, such studies will be critical to assess model performance. For generalizability, such future studies would need to be much larger, both in terms of the number of participants and the number of images and genetic conditions included.

Second, clinicians and non-clinicians exhibit different gaze behaviors when assessing images. This is not surprising, but quantifying these behaviors using methods like these may be helpful for activities such as ascertaining which phenotypic characteristics may be diagnostically important but which are frequently overlooked. Again, as AI support enters more and more clinical areas, these types of analyses may point to specific ways to augment the relationship between clinicians and AI tools. For example, data from extremely high-performing clinicians in human/classifier comparison experiments may be useful to design the education of less experienced clinicians or trainees, as well as AI tools.^9^

This study has several limitations. These include the number of participants and images viewed. The images also represent heterogeneous genetic conditions, and eye-tracking behavior may be affected by certain aspects of a particular image. While we grouped clinicians and non-clinicians into separate categories, there is varying experience and expertise within these groups, and differences in behavior between individuals. Additionally, our analyses focused on metrics like human visual attention, which may not reflect what is most important to a person evaluating an image. For example, an expert clinician may immediately perceive a key visual clue to a diagnosis and may then move on to spend more time observing less obvious features or searching the image for subtle clues. Overall, while our results show interesting trends, caution should be taken that they will generalize to other or all genetic conditions, to larger groups of individuals, or to other AI-based approaches.

As mentioned above, the results here point to multiple future possibilities, including involving larger numbers of participants and datasets to analyze further how well the results can be extrapolated. Related eye-tracking experiments could be used to explore multiple questions germane to genetics, such as with different data types (e.g., radiological studies or other physical examination features) encountered in clinical practice. Additional work could be done specifically quantifying different classifiers and saliency methods.

## Data Availability

Processed data produced in the present work are available in the manuscript and accompanying files, with code available via GitHub. Raw data are available upon reasonable request to the authors.

https://github.com/datduong/Tobii-AOI-FaceSyndromes

## Acknowledgements

This research was supported in part by the Intramural Research Program of the National Human Genome Research Institute, National Institutes of Health. This work utilized the computational resources of the NIH HPC Biowulf cluster.

