## Supplementary Materials for "Human and computer attention in assessing genetic conditions"

### Supplementary Material

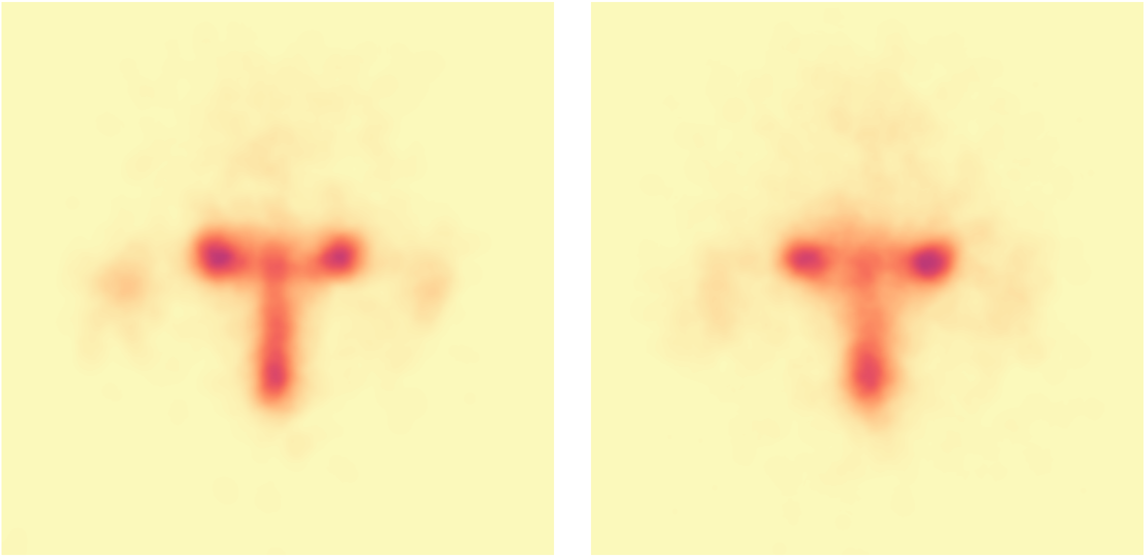

**Supplementary Figure 3**: We take the average of all the visual attention heat maps across all test images for the clinician (left) and non-clinician groups (right). We observed that most of the visual interests align with eyes, nose, and mouth areas. To account for normal human behavior when viewing an image, conditioned on the group expertise (clinician or non-clinician), we subtracted these common average areas from each individual heat map used for our analyses. This helps account for typical human behavior when viewing faces but does not cause us to ignore these areas of common facial attention.

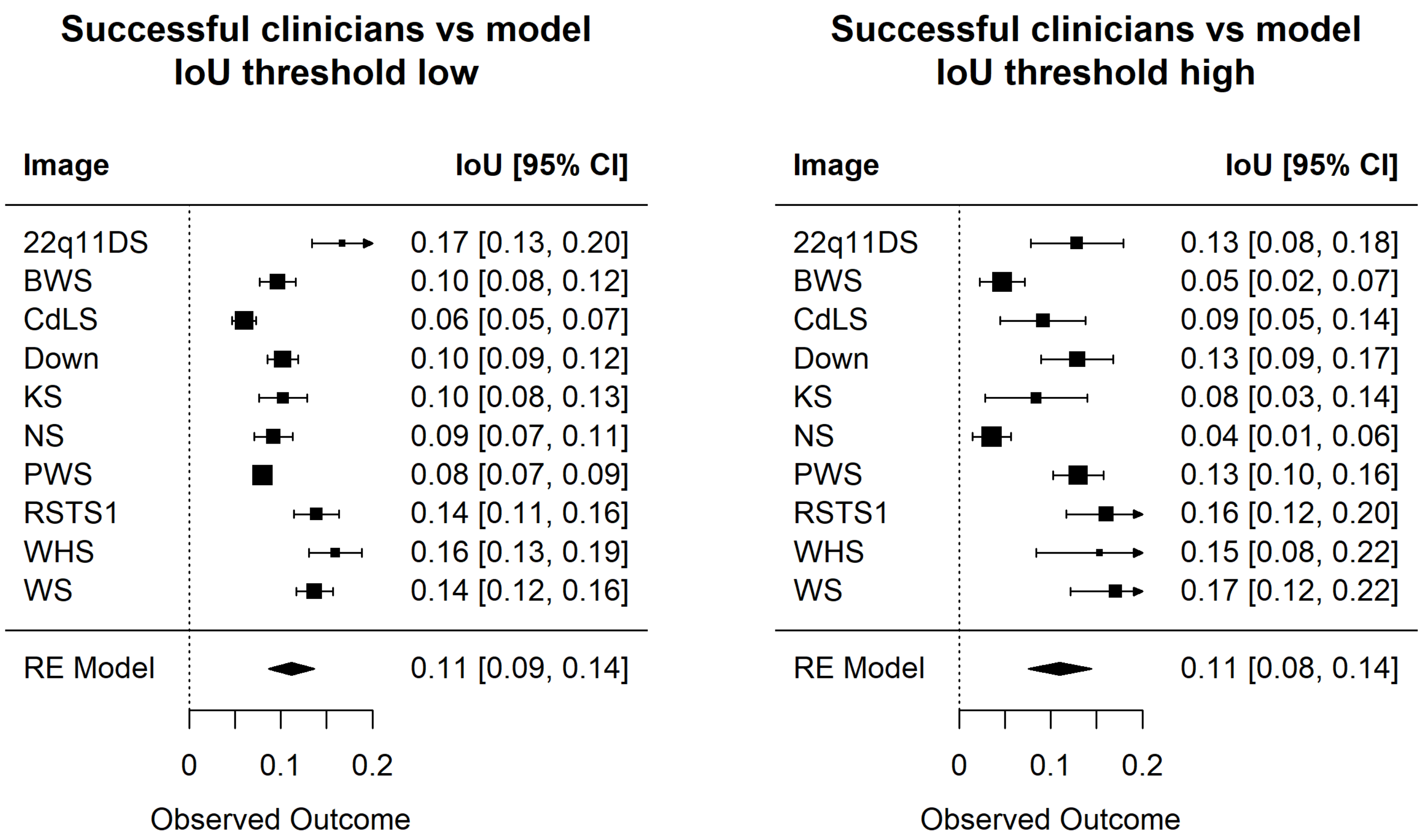


**Supplementary Figure 4**: IoU metric compares the visual attention of successful clinicians and model saliency maps over all the 10 test diseases at the same filtering threshold (e.g. low and then high thresholds for both saliency maps and Tobii heat maps).

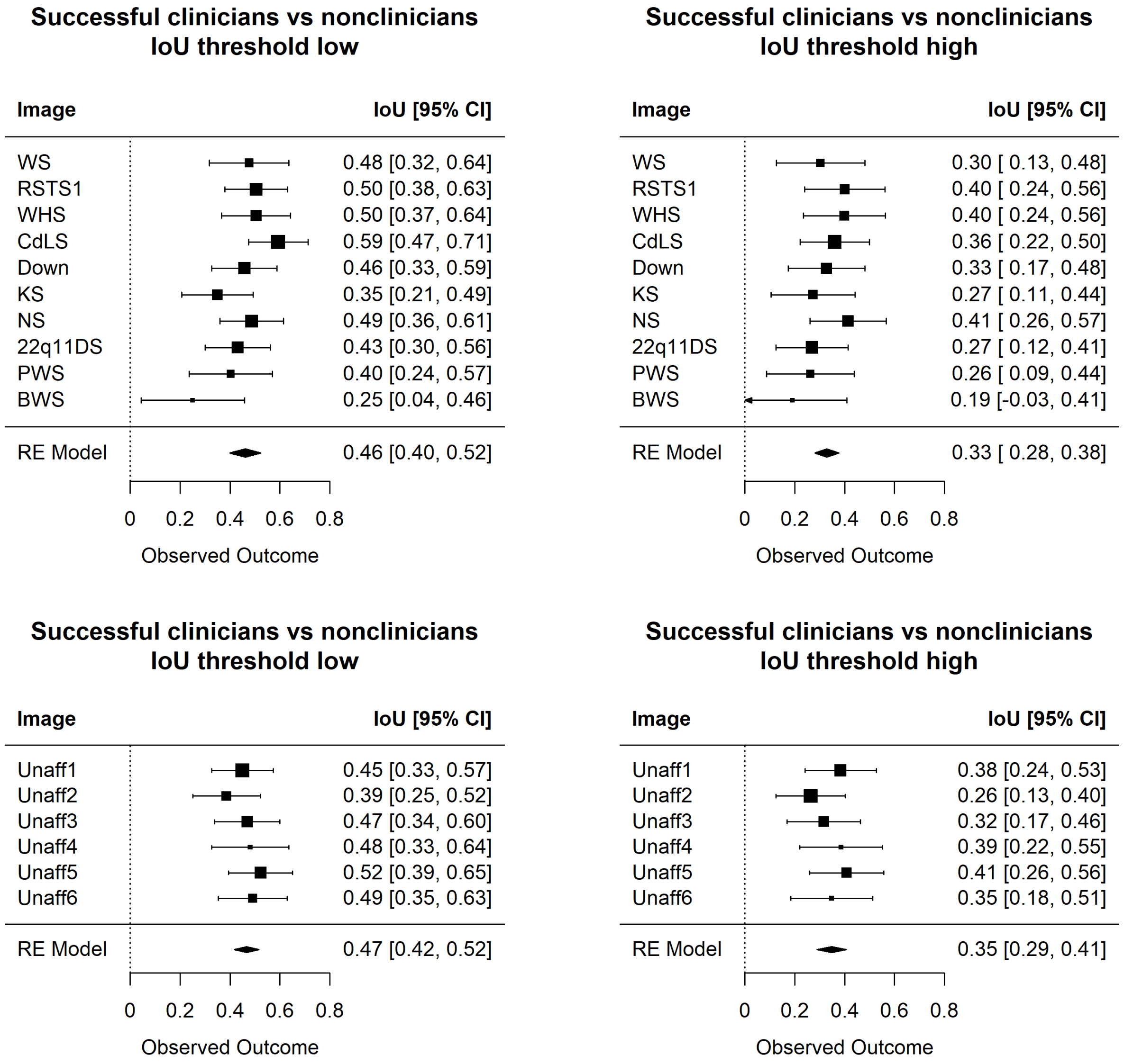


**Supplementary Figure 6**: IoU metric comparing the heat maps of successful clinicians versus non-clinicians conditioned on whether the test images are of affected or unaffected individuals.

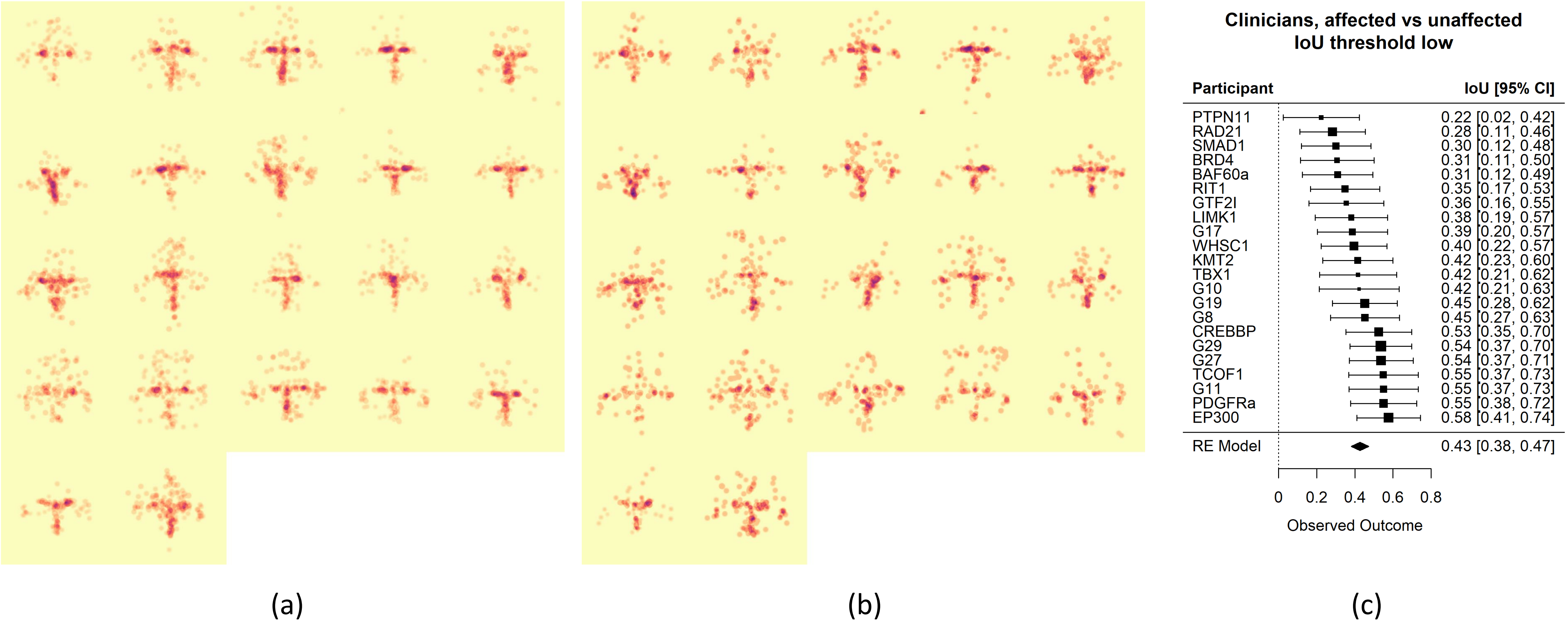


**Supplementary Figure 7**: For each of the 22 clinicians, we average heat maps over all 10 affected (a) and 6 unaffected (b) images. Each participant displays unique behavior. For example, in (a) the first participant (row 1 column 1) and sixth participant (row 2 column 1) showed unequal interest at the nose area. (c) We applied a low filtering threshold to remove spurious signals from the visual heat maps, and then estimated the differences in the visual attention between affected and unaffected images conditioned on the same participant. Across all participants, the average IoU score is far from zero, indicating that on average clinicians inspect similar facial features when classifying affected vs unaffected images.

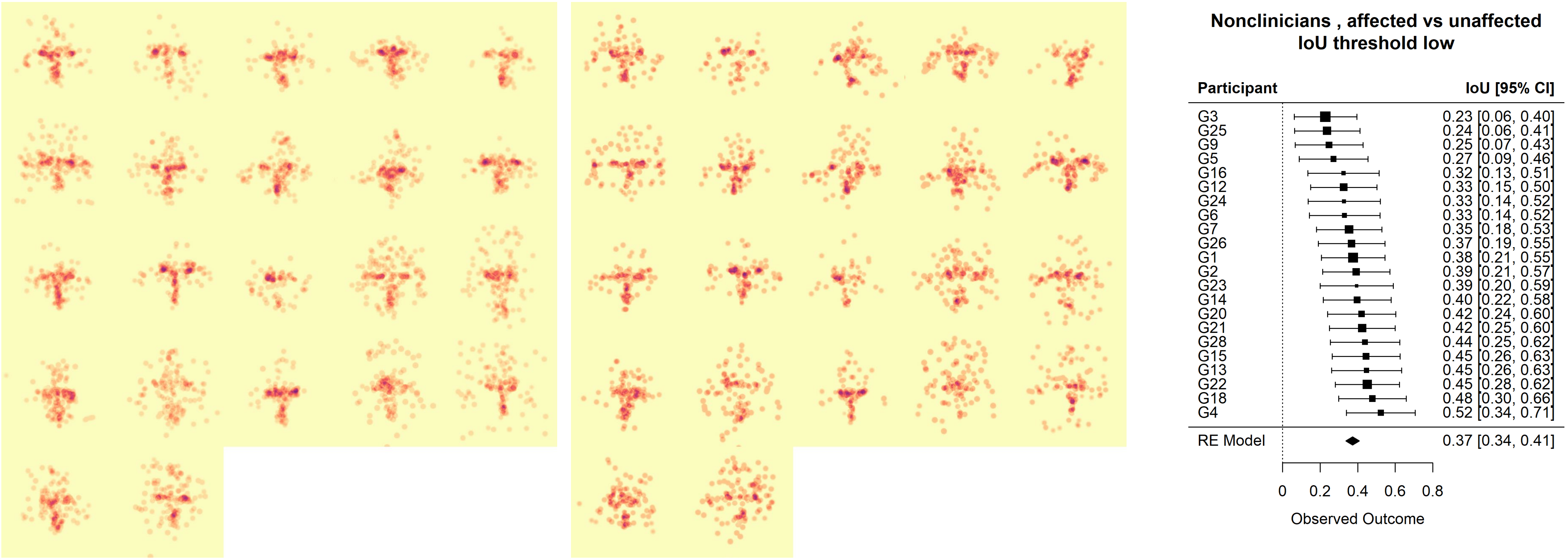


(a) (b) (c)

**Supplementary Figure 8**: For each of the 22 non-clinicians, we average heat maps over all 10 affected (a) and 6 unaffected (b) images. Each participant displays unique behavior; however, visually this uniqueness is less detectable compared to **Supplementary Figure 6**. We applied a low filtering threshold to remove spurious signals from the visual heat maps, and then estimated the differences in the visual attention between affected and unaffected images conditioned on the same participant. The average IoU score is far from zero, indicating that on average non-clinicians inspect similar facial features when classifying affected vs unaffected images.
